# Electoral processes and COVID-19 infections in Japan

**DOI:** 10.1101/2021.06.13.21258864

**Authors:** Shunsuke Murata, Takashi Nagata, Soshiro Ogata, Kunihiro Nishimura, Akihito Hagihara

## Abstract

Election campaigns and polling stations might be areas of potential infections; however, it is unknown whether elections can impact the spread of coronavirus disease (COVID-19). This study aimed to evaluate the association between election process and COVID-19 infection in Japan. Quasi-experimental design using time-series COVID-19 cases data with an election intervention. We chose the cities where elections for major and/or city/ward assembly members were held in 2021 January in Japan. The study period for each city spans one weeks before the start date of the election campaign and three weeks after the voting day (36 days). The daily number of patients testing positive for COVID-19. We used Poisson regression analysis with sandwich estimator to evaluate the association between election and the spread of COVID-19 infections. For Miyakojima City, we assessed the relationship between election and COVID-19 infections by using the Box-Jenkins autoregressive integrated moving average (ARIMA) model. We also estimated the instantaneous reproduction number (R_t_) in the city. There were 17 cities that met the inclusion criteria. In all models with three types of different lag effects (i.e., 4, 9, and 14 days), election was not a significant predictor of COVID-19 infections in the 17 cities. For Miyakojima City, the autoregressive integrated moving average (0, 0, 0) with a lag of 14 (α=14) was the best model. The partial coefficient (ω) of the election was 11.91 (95% confidence interval: 3.10–20.72, *P*<0.001), indicating that the election was associated with an increase in the number of COVID-19 cases 14 days after the election campaign (mean: 12 cases/day). R_t_ hovered far above 1 during and after the election campaign in Miyakojima. Although elections were associated with an increased number of COVID-19 cases in Miyakojima, this association was not verified in the analysis including all 17 cities. Therefore, if preventive measures prescribed by election guidelines are followed, elections do not necessarily relate to a spread of COVID-19 as long as the election process does not involve activities necessitating close contact.

## Introduction

Elections constitute an essential part of democracy.^1^ The ongoing coronavirus disease (COVID-19) pandemic poses a threat to people participating in the democratic process. In places scheduled to have elections, local authorities face the challenges of how and when to conduct polls, measures to reduce the COVID-19 cases associated with elections, and safety of election administration staff.^2,3^ An individual’s perceived risk of contracting COVID-19 and a city’s proximity to a COVID-19 cluster are associated with a lower likelihood of participation in voting.^4,5^

Campaign rallies and polling stations can be areas of potential infections, and voters may be reluctant to participate. There have been several reports on the association between elections and COVID-19 incidence. Specifically, according to studies on an election in Milwaukee, Wisconsin on April 7, 2020, there was no clear increase in infection cases, hospitalizations, or death after the election.^6,7^ However, in another study on a primary election on May 5, 2020 and COVID-19 infections in Michigan, Mississippi, and Missouri, mixed findings were reported. Although there was an association between voter turnout and COVID-19 infections in Michigan, no such association was observed in Mississippi or Missouri.^8^ A study on the national assembly elections in Korea reported that four voters who participated in the election tested positive for COVID-19.^9^

In the above elections conducted during the COVID-19 pandemic, election processes were held per guidelines published by the public authorities, such as the election commission, local government, Centers for Diseases Control and Prevention in the United States, and Korean Centers for Diseases Control in Korea.^6,9^ Thus far, findings on the association between public elections and the spread of COVID-19 are inconclusive. The number of elections included in the previous studies are limited.^6–9^ Thus, any generalizable conclusions from such studies are very limited. Moreover, it is not clear how those cities were chosen and if they are representative of the country as a whole. As such, it is very hard to interpret these results beyond the very local context of the cities. Moreover, whether the voters’ behaviors in the election process are conducive or detrimental to the prevention of COVID-19 infections are not fully clear. Therefore, in this study, we used all elections conducted during a prefixed period and evaluated the association between elections and COVID-19 infections in Japan.

## Methods

### Cities

We chose all cities where elections for mayor and/or city assembly members were held in January 2021 in Japan. Of the 24 cities which meet this criterion, there were 3 cities where Covid-19 case data could not be obtained (Towada, Ibaraki, and Iwade cities). There were no election campaigns in 4 cities (Usuki, Arao, Nomi, and Minamiawaji cities) because only one candidate ran for the mayoral election or the number of candidates for city assembly members was equal to vacancy. Thus, 17 cities were selected in this study. Those cities were Isesaki City, Kawagoe City, Toda City, Chiyoda Ward, Kikugawa City, Gotemba City, Iwakura City, Kameyama City, Takashima City, Unnan City, Kurashiki City, Kitakyushu City, Karatsu City, Yamaga City, Saito City, Nishinoomote City, and Miyakojima City in Japan (Fig. 1, Table 1).

**Table 1.** Summary of the elections in the 17 cities in Japan where mayoral and/or city assembly member elections were conducted in January 2021.

| City/ward* | Polling Day | Election type | Electorate | Voter | Voting rate | Population |
| --- | --- | --- | --- | --- | --- | --- |
| Isesaki | 2021/1/17 | mayoral election | 166,885 | 50,650 | 30.35 | 208,814 |
| Kawagoe | 2021/1/24 | mayoral election and city assembly members election | 290,275 | 64,008 | 22.05 | 350,745 |
| Toda | 2021/1/31 | city assembly members election | 108,606 | 42,225 | 38.88 | 136,150 |
| Chiyoda* | 2021/1/31 | mayoral election and ward assembly members election | 52,525 | 23,795 | 45.30 | 58,406 |
| Kikugawa | 2021/1/24 | mayoral election and city assembly members election | 37,016 | 22,227 | 60.05 | 46,763 |
| Gotemba | 2021/1/31 | mayoral election | 70,653 | 34,389 | 48.67 | 88,078 |
| Iwakura | 2021/1/24 | mayoral election | 38,061 | 12,538 | 32.94 | 47,562 |
| Kameyama | 2021/1/31 | mayoral election | 39,396 | 19,699 | 50.00 | 50,254 |
| Takashima | 2021/1/31 | mayoral elections and city assembly members election | 40,582 | 27,661 | 68.16 | 50,025 |
| Unnan | 2021/1/31 | mayoral election | 31,791 | 21,990 | 69.17 | 39,032 |
| Kurashiki | 2021/1/24 | city assembly members election | 393,400 | 139,173 | 35.38 | 477,118 |
| Kitakyushu | 2021/1/31 | city assembly members election | 787,960 | 317,472 | 40.29 | 961,286 |
| Karatsu | 2021/1/31 | mayoral election and city assembly members election | 98,976 | 56,888 | 57.48 | 122,785 |
| Yamaga | 2021/1/31 | mayoral election and city assembly members election | 42,896 | 29,309 | 68.33 | 52,264 |
| Saito | 2021/1/24 | mayoral election | 25,096 | 16,368 | 65.22 | 30,683 |
| Nishinoomote | 2021/1/31 | mayoral election and city assembly members election | 13,303 | 10,278 | 77.26 | 15,967 |
| Miyakojima | 2021/1/17 | mayoral election | 44,376 | 29,129 | 65.64 | 51,186 |

\*: Except for Chiyoda, every municipality is city. Ward and city are treated equally in the Public Offices Election Act (Law No. 100, 1950).

**Figure 1.**
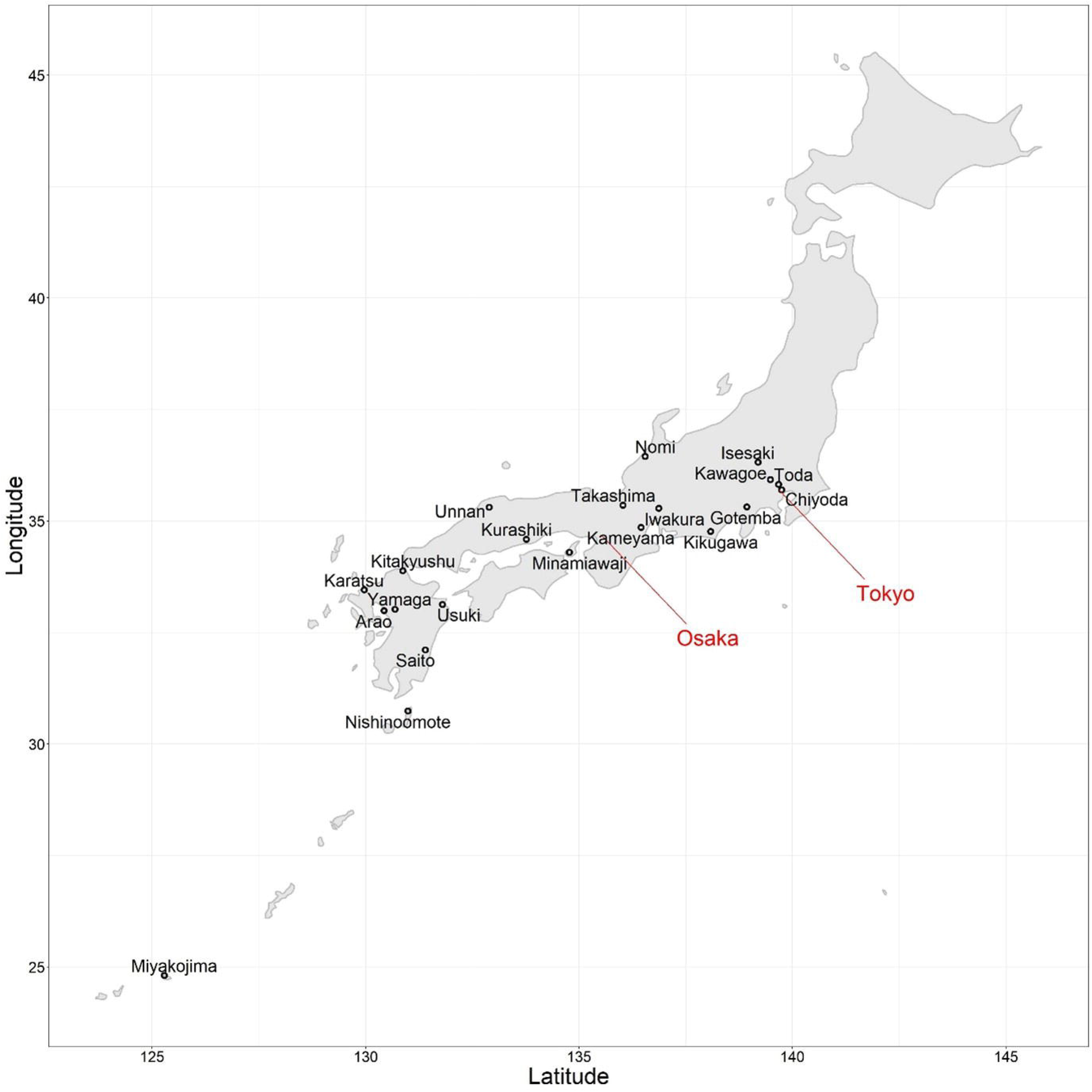
Location of the 17 cities in Japan. Brack points are locations of 17 cities used in this study. Red names (Tokyo and Osaka) are main prefectures in Japan.

### Elections

#### Election campaign

An election campaign is defined as activities to solicit electorates for balloting for the purpose of getting a specific candidate elected. The period of election campaign for mayor and city assembly members is 7 days after election announcement by the election commission of each city/ward, and activities are from 8 AM to 8 PM.^10^ Candidates are allowed to perform election campaigns using printed material, such as posters, flyers, postcards, newspaper advertisements, and the internet. Candidates can also perform election campaigns by speech, such as speech meeting, street oratory, calling a candidate name repeatedly on a campaign car, and political opinion broadcast.^10^ People other than candidates also can be engaged in election campaigns, such as delivering support speech at a gathering, asking their acquaintances for votes on the street, and soliciting others for votes by telephone and the internet.

#### Election guidelines during COVID-19 pandemic in Japan

In addition to the guidelines by the Public Offices Election Act (Law No. 100, 1950), special guidelines were issued for elections during the COVID-19 pandemic in Japan.^10–12^ Specifically, election campaigns and polling were conducted in accordance with the guidelines of the Ministry of Internal Affairs and Communication, Local Administration Bureau.^11^ For the election campaigns, considering a candidate’s freedom of political activities, each candidate was allowed to decide the mode of the campaign conducive to the prevention of virus infection. To prevent virus infections at the polling stations, early voting, increased number of polling stations, increased ventilation of voting rooms, mandatory wearing of face masks, physical distance at the station, and the use of voters’ private pens were encouraged.^11,12^

### Study period

The study period for each city spans two weeks before and three weeks after the voting date (36 days).

### Data collection

COVID-19 is categorized as a type 2 infectious disease by the law, and the daily number of positive COVID-19 cases (tested by polymerase chain reaction) have been reported by local authorities in Japan.^13^ Relevant data were obtained from the digital home pages of the 17 cities/ward (Supplementary Table 1). The study was approved by the institutional ethics committee of Kyushu university, and informed consent could not be obtained from the subjects since only aggregate data were used in the study.

### Data analysis

We used Poisson regression analysis with sandwich estimator to evaluate the association between elections and the spread of COVID-19 infections. The dependent variable was the number of patients with COVID-19 on the (t+α)^th^ day of the time series (Y_t+_ α). The independent variable was the election (“Election_t_”); “Election_t_” = 1 was set when t was in the election period, and “Election_t_” = 0 was set when t was outside the election period. As covariates, time (day 1 to day 36) were entered into the model. The natural log of the population was entered as an offset variable. The incubation period for COVID-19 is an average of 5–6 days or may be 14 days.^14^ Thus, we fitted the analysis model to daily COVID-19 case data when α takes the following numbers (i.e., 4, 9, 14).

We used the Box-Jenkins autoregressive integrated moving average (ARIMA) model to assess the relationship between elections and COVID-19 in Miyakojima. In view of the incubation period for COVID-19,^14^ we fitted the analysis model [Y_t+_ α = ARIMA (p, d, q) × (P, D, Q)_s_ + ωElection_t_] to daily COVID-19 case data by shifting α from 1 to 14 by 1, and selected the best-fit model based on Akaike’s Information Criterion. “Election_t_” = 1 was set when t was in the election period, and “Election_t_” = 0 was set when t was outside the election period. With respect to reported models, the stationarity of the time series data was verified by checking the plots of the autocorrelation functions and the roots of the polynomial (i.e., the size of the roots should be > 1).^15^ The model residuals were checked by autocorrelations at various lag times using the Ljung–Box chi-square statistic to confirm white noise.^15^

We also used a method by Cori et al.^16^ to estimate the instantaneous reproduction number (Rt) in Miyakojima, which is the average number of secondary cases generated by one primary case with the time of infection on day t, from 2 weeks before the start of the election campaign (i.e., January 10, 2021 for Miyakojima) through 3 weeks after the voting day (i.e., January 17, 2021 for Miyakojima). We assumed that the mean (standard deviation [SD]) of serial interval was 4.9 (±2.7).^17^ All *P*-values were two-sided, and statistical significance was set at *P*<0·05. All analyses were conducted using R (version 4.0.2; R Foundation for Statistical Computing, Vienna, Austria), and ARIMA model analyses were conducted using the “sandwich,” “forecast,” and “tseries” packages.

## Results

The population, electorates, and voting rates of the 17 cities are summarized in Table 1. Of the 17 cities, mayoral elections, city/ward assembly members elections, and elections for both mayor and city assembly members were held in seven, three, and seven cities/ward, respectively (Table 1). In every city, poling date was set on Sunday of January 17, 24, or 31. Regarding our internet search, elections in the 16 cities except for Miyakojima City were conducted following the relevant election guidelines.^10–12^ Daily COVID-19 case counts during the study period (36 days) in the 17 cities are summarized in Table 2, and COVID-19 case counts per 100,000 persons in the 17 cities are shown in Figure 2. The median COVID-19 case counts in the 17 cities during the study period ranges from 0 to 15.5. When COVID-19 case counts are calculated per 100,000 persons, the adjusted case counts of 16 cities were very similar except for Miyakojima city. In Miyakojima, there were 266 COVID-19 cases; the mean count (standard deviation [SD]) was 7.39 (±10.13), with a minimum and maximum count of 0 and 57, respectively. Table 3 shows the results of the Poisson regression analyses of the mayoral and/or city assembly elections and COVID-19 infections in the 17 cities. In all of the models with three types of different lag effects, the election was not a significant predictor. Specifically, the risk ratio of the election was 1.00 (0.66–1.51) in a model with 14 days lag effects, 0.94 (0.66–1.32) in a model with 9 days lag effects and 1.02 (0.79–1.33) in a model with a lag effect of 4 days.

**Table 2.**
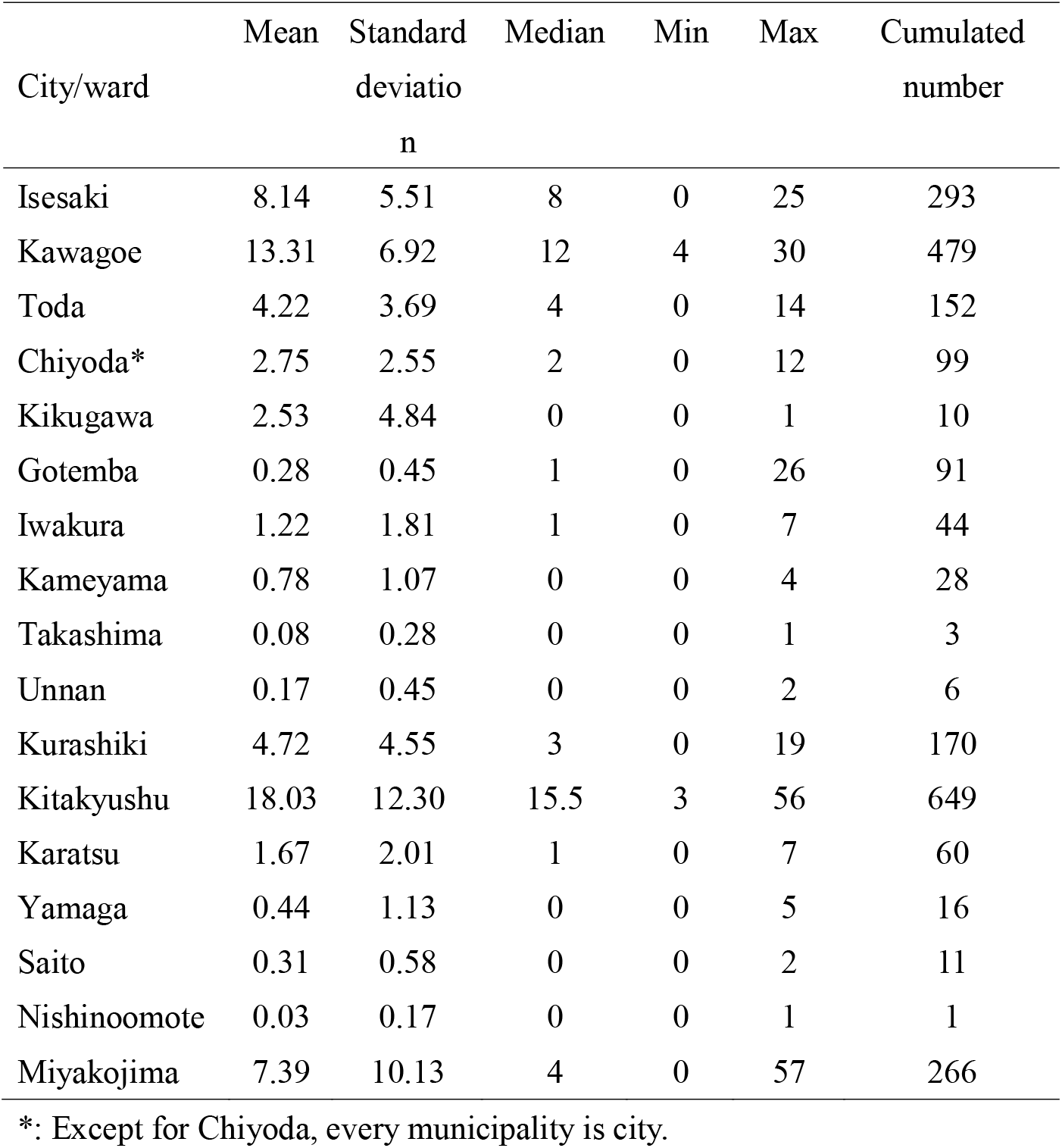
Summary of COVID-19 infections in the 17 cities during 36 days of the study period.

**Table 3.** The association between election processes and COVID-19 infections in the 17 cities with three different lag effects.

|  | Risk ratio (95% confidence interval) |
| --- | --- |
| Election with 14 days lag ($\alpha$) | 1.00 (0.66–1.51) |
| Election with 9 days lag ($\alpha$) | 0.94 (0.66–1.32) |
| Election with 4 days lag ($\alpha$) | 1.02 (0.79–1.33) |

Note: The results of Poisson regression analyses with sandwich estimator. The dependent variable ( $Y_{t+\alpha}$ ) is the number of patients with COVID-19 on the $(t+\alpha)^{\text{th}}$ day. The independent variable ( $X_t$ ) is election (“Election<sub>t</sub>”). Time (day 1 to day 36) and log of population as offset variable are also entered into the model.

**Figure 2.**
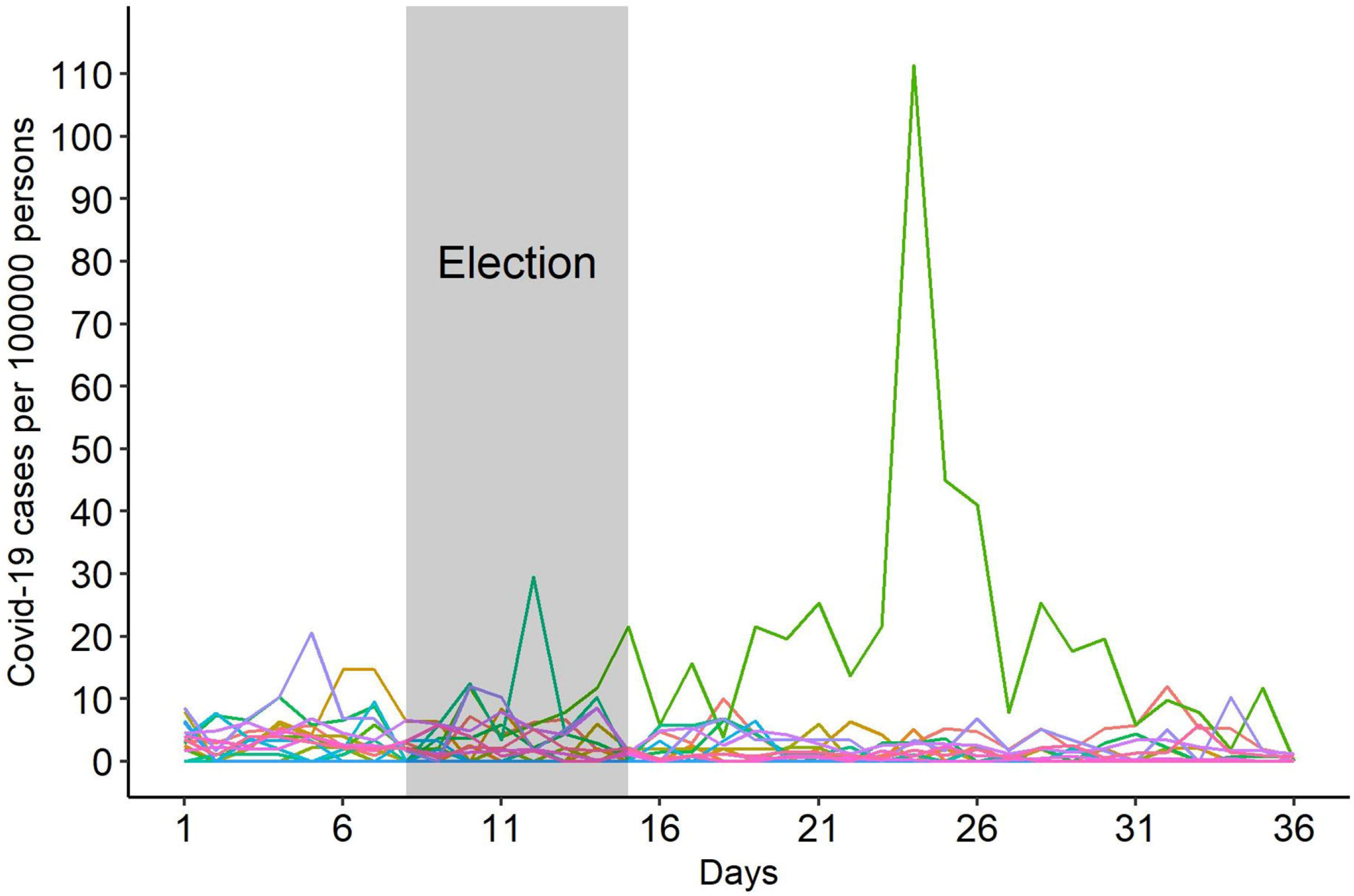
Changes in the number of COVID-19 cases during the study period in the 17 cities. Each line indicates the daily number of cases per 100000 persons in each city, and the green line indicates a change in the number cases in Miyakojima City.

COVID-19 case counts per 100,000 persons in Miyakojima was much larger than counts in the rest of the 16 cities. Thus, we used the Box-Jenkins autoregressive integrated moving average (ARIMA) model to assess the relationship between elections and COVID-19 in Miyakojima City. Table 4 shows the results of the ARIMA regression analyses of the mayoral election and COVID-19 infection. Of all models shifting α from 1 to 14 by 1, a model with a lag of 14 was the best-fit model based on Akaike’s Information Criterion. In the table, models with 4 and 9 days of lag are also reported for reference. ARIMA (0, 0, 0) with a lag of 14 (α=14) was the best model. The Ljung–Box test statistic was 2.39 (*P*=0.496). These statistics indicate the adequacy of the model. The partial coefficient (ω) of the election was 11.91 (95% confidence interval: 3.10 to 20.72, *P*<0.001), which implies that elections were associated with an increase in the number of COVID-19 cases at 14 days after the election campaign by an average of 12 cases per day. As for ARIMA (1, 0, 0) with a lag of 4 (α=4), the Ljung–Box test statistic was 3.06 (*P*=0.383), and the unit-root of AR (1) was 2.70. Although these statistics indicate the adequacy of the model, the partial coefficient (ω) of the election was −1.68 (95% confidence interval: −12.32 to 8.96) and insignificant. As for ARIMA (1, 0, 0) with a lag of 9 (α=9), the Ljung–Box test statistic was 2.73 (*P*=0.436). The unit-root of AR (1) was 2.65. Although these statistics also indicate the adequacy of the model, the partial coefficient (ω) of the election was 10.97 (95% confidence interval: −0.93 to 22.86) and insignificant. Figure 3 shows the instantaneous reproduction number (R_t_) during the 6 weeks including 1 week of the election campaign in Miyakojima. The reproduction number (R_t_) hovered far above 1 from before the start of the election campaign through 2 weeks after the voting day.

**Table 4.**
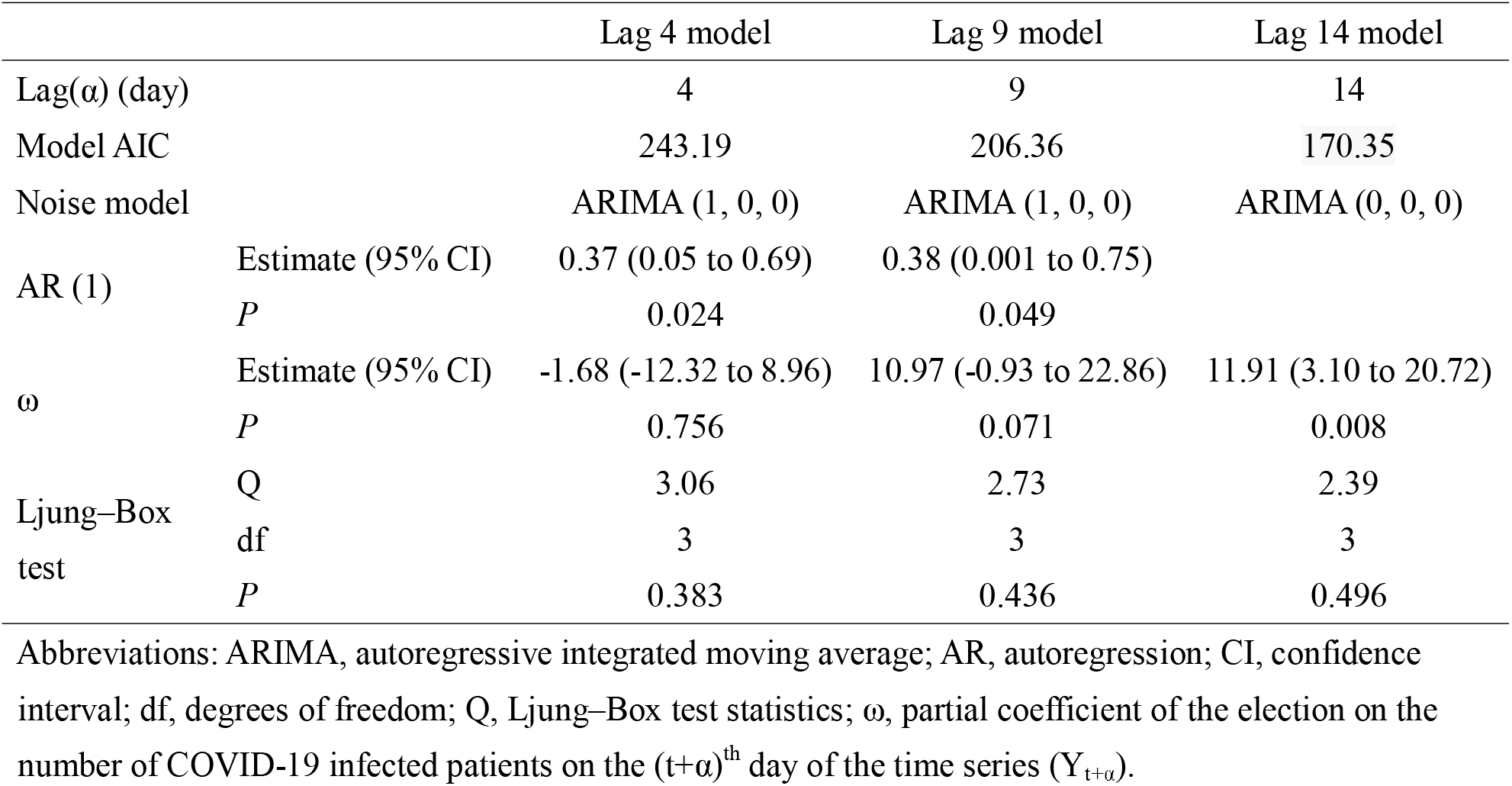
Association between election and coronavirus infections in Miyakojima city by ARIMA regression analyses with different lags.

**Figure 3.**
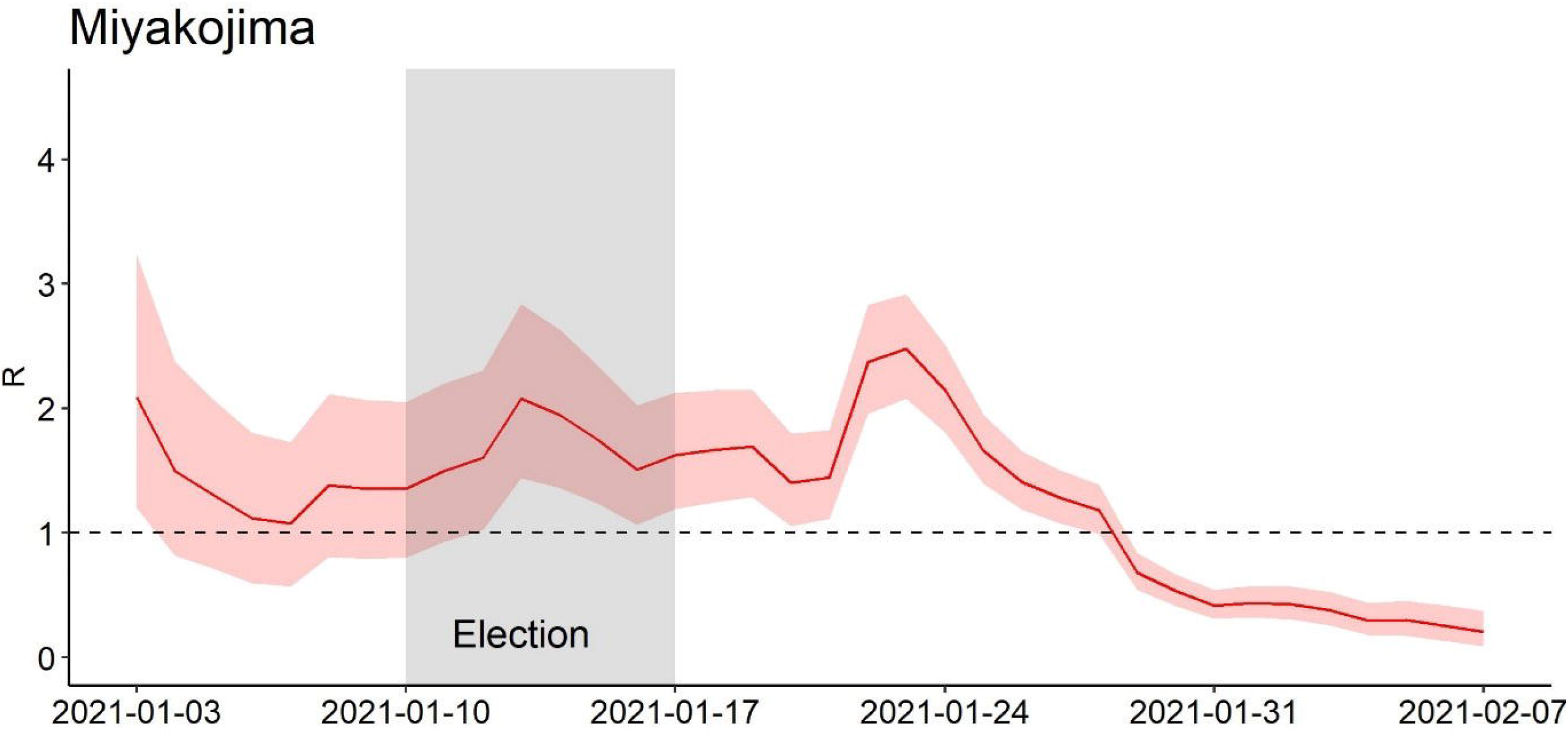
Estimated instantaneous reproduction number R_t_ from two weeks prior to the start of election campaign through three weeks after the voting day in Miyakojima City. Red line indicates reproduction number R_t_, and shaded areas indicates the 95% confidence interval.

## Discussion

An analysis of the association between elections and COVID-19 cases showed mixed results. Specifically, an association between elections and subsequent COVID-19 cases was not verified in the 17 cities. However, an increase in COVID-19 cases was observed after the election in Miyakojima City. The reproduction number before and after the election campaign was also well above 1 after the election in Miyakojima. The reason for the difference in these findings might be victory celebrations by the mayor-elect group, and consolation gatherings by defeated candidates held on January 17, 2021, in Miyakojima. It has also been reported that a large number of gatherings with facilities for eating and drinking were included in the campaigns in Miyakojima.^18,19^ This is consistent with the increasing production numbers well over 1 during the election campaign period in Miyakojima. However, there were no such incidents reported in the rest of 16 cities. Among the 17 cities, income level as a surrogate of health literacy was lower, and the voting rate was higher in Miyakojima. These factors might have also contributed to the differences in our results. Another explanation is the geographical difference between Miyakojima Island and the rest of the cities as urban areas. Considering mobility, island settings might cause more frequent contact than urban areas, resulting in COVID-19 infection.

There are some practical implications of our findings in preventing virus infections in the election process. Although an increase in COVID-19 cases was observed after the election in Miyakojima City, an association between elections and subsequent COVID-19 cases was not verified in the analyses based on the 17 cities. Voting rate reflects voters’ interest in election. Although there were four elections in Kameyama, Unnan, Yamaga, and Nishinoomote cities that showed higher voting rates than that in Miyakojima (i.e., 65.64%) after January 17, 2021 (Table 1), there was no report concerning gatherings with facilities for eating and drinking in those cities. In view of these findings, government guidelines may be effective in preventing COVID-19 infections in elections so far as election campaigns including gatherings with facilities for eating and drinking are avoided.^11^ Our findings are also consistent with previous research.^2,3^ In this ecological study, all elections for city assembly members and mayors during fixed periods (i.e., January 2021) in Japan were evaluated. Thus, the findings are conclusive, and the results of the study are of scientific value due to the wide scope of the study. Since all elections are studied in a fixed period, we believe that the findings are generalizable to other areas with similar SES and political and election system.

Additionally, there were several notable findings in the study. First, instantaneous reproduction number hovered at 1 before the election campaign in Miyakojima. Although election campaign prior to the election announcement by the election commission is not allowed, political activities such as advertising a political party’s policy and gaining more seats are allowed.^10^ It has been reported that pre-election campaigns with drinking and eating opportunities, such as joint speech meetings by candidates, election support and campaign rally, were held before election announcements in Miyakojima city.^19^ The high instantaneous reproduction number before the election campaign might be due to pre-election campaign in Miyakojima. Second, an association between elections and subsequent COVID-19 cases was not verified in Nishinoomote, an island city (Table 2, Figure 2). Since COVID-19 infection cases after the election in Miyakojima was reported,^18,19^ election campaigns resulting in close contact, such as campaigns with drinking and eating opportunities, have been suggested to be a cause of increased number of COVID-19 infections in Miyakojima.^20^ Thus, election commissions issued warnings against election campaigns that might result in close contact after January 17, 2021 (*i*.*e*., voting date in Miyakojima). There was no report on this type of election campaign in Nishinoomote city either. Elections in accordance with the guideline might be attributable to the finding of no association between election and COVID-19 infections in Nishinoomote.^11,12^ Third, the incubation period for COVID-19 is wide, with an average of 5–6 days and may as long as 14 days.^14^ In the analysis of Miyakojima, we firstly revealed that ARIMA regression with 14 days of lag was the best-fit model based on Akaike’s Information Criterion. It was implied that the longest lag of 14 days is applicable to population data on COVID-19 infections.

Our study has a few limitations. First, a causal link between elections and COVID-19 cases could not be determined. Second, while we believe our findings are conclusive and has some external validity, the findings are not applicable to countries with different socio-economic statuses or COVID-19 incidence rates.

In summary, our data show that elections conducted with adequate preventive measures for election campaigns and voting do not necessarily relate to an increased number of COVID-19 cases in areas with similar socio-economic statuses and levels of infection, if activities resulting in close contact are not involved in the election process.

## Data Availability

Access to data and the R codes are available from the corresponding author on request.

## Acknowledgement

We are grateful to Ms. Naoko Hagihara and Ms. Yuka Yoshida for their assistance in collecting relevant literature and the COVID-19 infections data for our study. The authors have no conflicts of interest to declare. There was no role of the funding source in the study.

## Notes

### Competing Interest Statement

The authors have declared no competing interest.

### Author Declarations

The study was approved by the institutional ethics committee of Kyushu university, and informed consent could not be obtained from the subjects since only aggregate data were used in the study.

