## Supplemental table 1 for "Electoral processes and COVID-19 infections in Japan"

| Supplemental table 1. Digital home pages of the 17 cities/wards from where relevant data are retrieved. | |
| --- | --- |
| City/ward | URL |
| Isesaki | http://stopcovid19.pref.gunma.jp/ |
| Kawagoe | https://opendata.pref.saitama.lg.jp/data/dataset/covid19-jokyo |
| Toda | https://opendata.pref.saitama.lg.jp/data/dataset/covid19-jokyo |
| Chiyoda* | https://catalog.data.metro.tokyo.lg.jp/dataset/t000010d0000000085 |
| Kikugawa | https://opendata.pref.shizuoka.jp/dataset/8167.html |
| Gotemba | https://opendata.pref.shizuoka.jp/dataset/8167.html |
| Iwakura | https://www.city.iwakura.aichi.jp/0000004636.html |
| Kameyama | https://www.pref.mie.lg.jp/YAKUMUS/HP/m0068000066_00002.htm |
| Takashima | http://www.city.takashima.lg.jp/www/contents/1593163379414/index.html |
| Unnan | https://shimane-covid19.com/ |
| Kurashiki | https://okayama.stopcovid19.jp/ |
| Kitakyushu | https://ckan.open-governmentdata.org/dataset/8a9688c2-7b9f-4347-ad6e-de3b339ef740 |
| Karatsu | https://stopcovid19.code4saga.org/ |
| Yamaga | https://www.pref.kumamoto.jp/soshiki/211/82808.html |
| Saito | https://www.pref.miyazaki.lg.jp/fukushihoken/covid-19/kenmin/20200923170835.html |
| Nishinoomote | https://covid19.code4kagoshima.org/ |
| Miyakojima | https://www.pref.okinawa.lg.jp/site/hoken/kansen/soumu/press/20200214_covid19_pr1.html |

*: Except for Chiyoda, every municipality is city.
